# Time to publication in medical education journals: An analysis of publication timelines during COVID-19 (2019-2022)

**DOI:** 10.1101/2024.01.04.24300847

**Authors:** L.A. Maggio, J.A. Costello, K. R. Brown, A.R. Artino, S.J Durning, T. Ma

## Abstract

**Introduction:** COVID-19 changed scholarly publishing. Yet, its impact on medical education publishing is unstudied. Because journal articles and their corresponding publication timelines can influence academic success, the field needs updated publication timelines to set evidence-based expectations for academic productivity. This study attempts to answer the following research questions: did publication timelines significantly change around the time of COVID-19 and, if so, how?

**Methods:** We conducted a bibliometric study; our sample included articles published between January 2018, and December 2022, that appeared in the Medical Education Journals List-24 (MEJ-24). We clustered articles into three time-based groups (pre-COVID, COVID-overlap, and COVID-endemic), and two subject-based groups (about COVID-19 and not about COVID-19). We downloaded each article’s metadata from the National Library of Medicine and analyzed data using descriptive statistics, analysis of variance, and post-hoc tests to compare mean time differences across groups.

**Results:** Overall, time to publish averaged 300.8 days (*SD* = 200.8). One-way between-groups ANOVA showed significant differences between the three time-based groups *F* (2, 7473) = 2150.7, *p* <.001. The post-hoc comparisons indicated that COVID-overlap articles took significantly longer (*n* = 1470, *M*= 539; *SD* = 210.6) as compared to pre-COVID (*n* = 1281; *M* = 302; *SD* = 172.5) and COVID-endemic articles (*n* = 4725; *M* = 226; *SD* = 136.5). Notably, COVID-endemic articles were published in significantly less time than pre-pandemic articles, *p* < .001.

**Discussion:** Longer publication time was most pronounced for COVID-overlap articles. Publication timelines for COVID-endemic articles have shortened. Future research should explore how the shift in publication timelines has shaped medical education scholarship.

## Introduction

Journal articles are an important metric of academic productivity, and publication success is a significant consideration in tenure and promotion decisions. However, the publication process and its timeliness are influenced by factors beyond the quality of the scholarship (e.g., reviewer availability, quality, and responsiveness). Prior to the COVID-19 pandemic it took, on average, 263 days from submission of an article to a medical education journal for it to appear in PubMed [1]. This finding was based on articles published between 2008-2018. Since this publication, the COVID-19 pandemic has changed the world and consequently scholarly communication [2]. While multiple studies have investigated the impact of COVID-19 on publishing broadly [3,4] and in biomedicine more specifically [5], the pandemic’s effect on medical education publishing is unstudied.

COVID-19 was a “game changer” in medical education [6]. Yet, it is unknown how COVID-19 shaped publishing in medical education and if publication-related factors observed in other fields (e.g., publication type and coverage of COVID-19) played a role in publication timelines. As publications and their corresponding timelines are crucial determinants of academic success, it is critical for the field to have an updated publication timeline. Accurate timelines enable stakeholders to set realistic expectations for productivity and, as needed, to advocate for their members. Publication timelines also provide the consumers of these publications with a sense of the recency of an article’s content, which is important because findings from such articles can often influence educational policy and practice.

COVID-19 shifted the dynamics of scholarly publishing. Researchers found that on the one hand, COVID-19 “turbocharged” some scholarly publishing [5] such that the speed and volume of publication increased to provide rapid evidence to front-line healthcare workers, policy makers, and the public [3,7]. For example, one study found that early in the pandemic, medical journals halved their time to publication [8] and articles about the pandemic itself were published at faster rates and higher volumes than those not related to the pandemic [2,9,10]. On the other hand, publication of non-COVID focused research slowed [2], the productivity of certain groups (e.g., females, physician scientist trainees, caregivers, medical faculty) diminished [11–15], and the lack of access to classrooms and labs delayed research that was in the data collection and analysis phase.

Journal publishers and editors responded to COVID-19 in a variety of unprecedented ways. For example, to expedite publication, 20 major publishers and scholarly organizations united to establish the C19 Rapid Review initiative through which they shared peer reviews and the identities of reviewers thus streamlining the flow of manuscripts across journals [16]. In two other instances, the journal *eLife* adapted its peer review policies such that reviewers were advised not to require additional experiments or analyses in revision requests, because these requests would hinder publication speed [17], and the *Lancet* expanded their ‘fast tracking’ system for articles to expedite the sharing of ‘critical knowledge’ [5]. Recognizing the pandemic’s toll on reviewers and authors, many journals, including those in medical education, extended peer review and revision timelines [18]. Across science broadly there is some evidence as to how such initiatives impacted publication timelines [19,20], but there is no evidence specific to medical education.

At the start of the pandemic, *eLife’s* editor-in-chief espoused: “Publishing will not and should not be anybody’s first priority in the coming months” [17]. However, now that we are well beyond the pandemic’s emergency period, it is hard to ignore academia’s long history of prioritizing journal articles as a marker of success [21], which has high stakes implications for the scholarly progression of individuals and the field. At the individual level, journal articles and their related publication timelines play a role in one’s ability to be promoted, to secure grant funding, and to graduate from publication-dependent degree programs. For example, a review of promotion and tenure guidelines for faculty of medicine at 92 universities identified that 95% of guidelines mentioned peer-reviewed publications with 35 institutions (38%) specifying a required number of articles [22]. Thus, it is important for individuals and administrators to have updated publication timelines that reflect the reality of publishing during COVID-19.

Although the pandemic has ended and COVID-19 is now endemic, the potential for future pandemics or natural disasters is high, such that understanding these timelines is still an important aim. First, it will help to ensure that realistic expectations for productivity are being set and that evidence is available for advocating for researchers who may have faced a publication timeline influenced by the pandemic. An updated publication timeline also informs evidence-based decision making. Ideally, individuals make decisions using the “best available evidence” [23] with the currency of that evidence being a factor that enables the user to determine the relevance of the evidence to their current context. Thus, in this study, we explore publication timelines in medical education just before and during the COVID-19 pandemic. To guide this study, we asked: did publication timelines significantly change around COVID-19 and, if so, what were those changes, both broadly and in relation to publishing characteristics such as publication type and whether or not an article was about COVID-19.

## Methods

We conducted a bibliometric study that replicated and expanded upon a prior study on publication timelines in medical education [1]. That prior work drew heavily upon an earlier study that had investigated this topic broadly across scientific disciplines [24]. Where deviations were made from the original study [1] we make note and provide a rationale.

### Sample

We included articles that appeared in the Medical Education Journals List (MEJ-24) and that are indexed by PubMed. The MEJ-24 is a seed set of journals that was derived using bibliometric methods to represent the field of medical education [25]. The MEJ-24 includes 24 journals and represents an expansion from the previous study, which considered 14 journals [1].

Our sample included articles published in any one of the MEJ-24 journals between January 1, 2018 and December 31, 2022. To investigate the potential differences in publication times before and after the COVID-19 pandemic, we clustered articles into the following three groups (See Figure 1). The creation of these three groups deviates from the earlier study, which considered all articles across the study period at the same.

**Figure 1:**
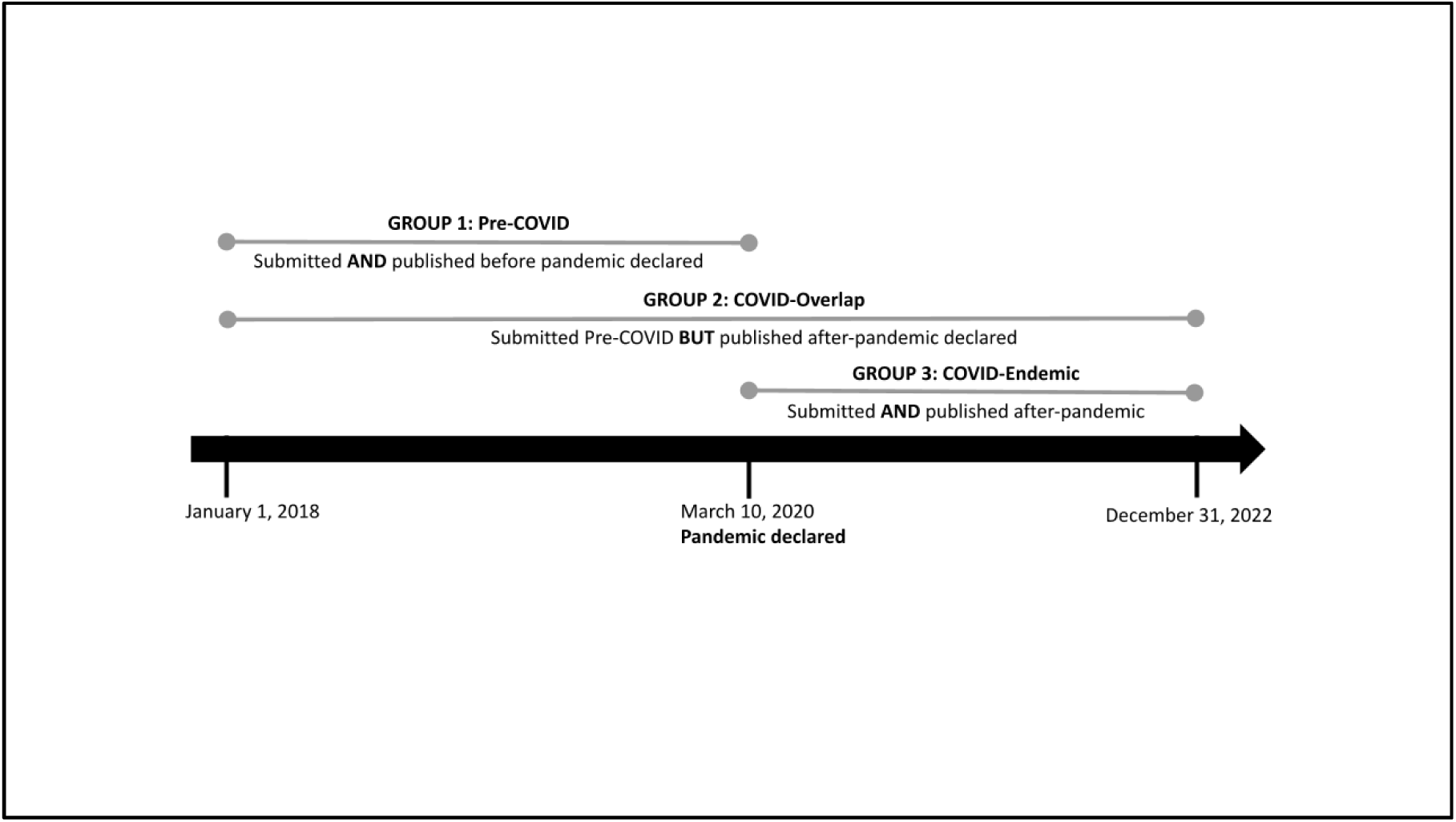
Pre-COVID, COVID-overlap, and COVID-endemic periods.

Group 1 (G1; pre-COVID) included articles submitted between January 1, 2018–March 10, 2020 and published prior to March 10, 2020. We selected March 10, 2020 as a cut off date for publication as the World Health Organization declared a global pandemic on this date [26]. This group served as our control, since the COVID-19 pandemic should not have affected these articles’ publication time. We conceptualized Group 2 (G2; COVID-overlap) as an overlap period which contains articles submitted before the COVID outbreak, but not published until after the COVID outbreak. G2 articles were submitted between January 1, 2018 and March 10, 2020 and published after March 10, 2020. Group 3 (G3; COVID-endemic) included articles submitted after March 10, 2020 and published by December 31, 2022. We descriptively summarize the number of days in publication timeline for these three periods and compare the mean-level differences for statistical significance.

### Data Collection

To collect our data, we referenced *The History of Publication Delays* [27] an analysis of publication delays up to 2015 utilizing the entire PubMed database. We used the original code created by Himmelstein [28] that is deposited on his Github account with only minor modifications. The modifications included an update for a depreciated function within a Python package and minor adjustments to account for a smaller dataset. On October 10, 2023 JC queried the eSearch e-utility, an API interface, using the updated code to download summary metadata from the National Library of Medicine (NLM) for all included articles [29]. Metadata included: Journal name, PubMed ID (PMID), NLM Journal Id, Article Type, Citation, Abstract, References, and Article History (Received Date, Accepted Date, and PubMed Publication Date). To be included in the analysis, it was necessary that an article was received by a publisher in the MEJ-24 on or after January 1, 2018 and published by the same journal before December 31, 2022. All metadata was merged as a table and exported to a CSV file.

We relied on the NLM’s indexing to determine an article’s publication type (e.g., review, letter, clinical trial) and if it was about COVID-19. To determine publication types, we used the publication types as defined and applied by the NLM. To characterize articles about COVID, we utilized NLM LitCOVID, which is a comprehensive search of PubMed for articles about COVID [30,31]. We made the judgment that an article was likely not peer reviewed, by identifying the articles with the following NLM publication types: Comment, Editorial, Letter, News, Published Erratum, or Retracted Publication. All other publication types were considered to be peer reviewed.

We also needed to account for articles where the publisher submitted an accepted date that was prior to a received date (*n* = 18) or a published date that is prior to an accepted date (*n* = 5).

### Analysis

Data were analyzed using SPSS version 28 [32]. To analyze the data, we first reported descriptive statistics. To compare publication timeline differences for articles that were published pre-COVID and after-COVID (the latter is further separated into articles submitted pre-COVID and after-COVID), we conducted analyses of variance (ANOVA) and compared mean differences across groups. Upon the significance level of an ANOVA Omnibus *F* test (*p* < .05), a follow-up LSD post hoc pairwise comparison was conducted to determine where the significance was between any one of the three group comparisons. We also conducted independent-samples t-tests to compare mean differences in publication timeline between COVID vs. non-COVID related articles. Timeline differences in articles of different publication types were also explored. Cohen’s *d* was reported upon significance to determine the effect sizes.

## Results

During the study period 16,544 articles were published in MEJ-24 journals (See Zenodo for dataset [33]). Of these journals, 15 supplied partial or complete publication timeline data representing 8,501 (51.4%) articles (See Online Appendix A). For the purposes of our research question, we focused on articles submitted after January 1, 2018 and published prior to December 31, 2022 with full timeline data. This represented 7,614 articles from 15 journals of which 23 articles included flawed metadata (e.g. the received date was after the acceptance date) so were removed resulting in 7,591 (45.9%) articles included for analysis.

The time to publish was on average 188.1 days (*SD* = 115.3). Time from submission to acceptance was on average 149.9 days (*SD* = 106.5). Processing time, which is the time between acceptance and appearing in PubMed, was on average 38.2 days (*SD* = 46.1).

One-way between-group ANOVA results showed the publication time was significantly different among Group 1 (pre-COVID), Group 2 (COVID-overlap), and Group 3 (COVID-endemic), *F* (2, 7588) = 237.66, *p* <.001. Significant differences were also found for acceptance time, *F* (2, 7588) = 177.10, *p* <.001, and process time, *F* (2, 7588) = 79.58, *p* <.001. These results indicated publication timeline was significantly different during pre-COVID (G1), COVID-endemic (G3), and when there is overlap during COVID (G2, i.e., submitted pre-COVID but published after-COVID).

The post-hoc comparisons using the Tukey HSD test indicated that articles of Group 2 (COVID-overlap, *n* = 761, *M*= 262.8; *SD* = 134.4) took significantly longer time in publication as compared to articles in Group 1 (both submitted and published before COVID, *n* = 2004; *M*= 200.7; *SD* = 118), and those in Group 3 (both submitted and published COVID-endemic, *n* = 4826; *M*= 171.1; *SD* = 105.2). Significant results with the same trend were found for acceptance time, and for processing time (see Table 1 for *M*s and *SD*s).

**Table 1.** Publication timelines for articles published in the MEJ-24 between 2018-2022 with available publication metadata.

|  | N | Time to Publication |  |  | Time to Acceptance |  |  | Processing Time |  |  |
| --- | --- | --- | --- | --- | --- | --- | --- | --- | --- | --- |
|  |  | Median | Mean | SD | Median | Mean | SD | Median | Mean | SD |
| Overall articles | 7591 | 165.0 | 188.1 | 115.3 | 129.0 | 149.9 | 106.5 | 23.0 | 38.2 | 46.1 |
| *Time Frame* |  |  |  |  |  |  |  |  |  |  |
| G1-Pre-COVID | 2004 | 181.0 | 200.7 | 118.0 | 133.0 | 154.7 | 109.0 | 27.0 | 46.0 | 53.3 |
| G2-COVID-overlap | 761 | 246.0 | 262.8 | 134.4 | 197.0 | 213.6 | 129.4 | 24.0 | 49.1 | 57.3 |
| G3-COVID-endemic | 4826 | 151.0 | 171.1 | 105.2 | 119.0 | 137.9 | 97.3 | 23.0 | 33.3 | 39.7 |
| **COVID-Related** |  |  |  |  |  |  |  |  |  |  |
| Non-COVID-related | 6736 | 169.0 | 192.1 | 116.7 | 133.0 | 153.5 | 107.8 | 24.0 | 38.5 | 46.3 |
| COVID-related | 855 | 141.0 | 156.8 | 98.5 | 99.0 | 121.1 | 90.6 | 22.0 | 35.7 | 44.3 |
| G3-Non-COVID-related | 3975 | 152.0 | 174.2 | 106.5 | 123.0 | 141.4 | 98.4 | 23.0 | 32.8 | 38.7 |
| G3-COVID-related | 851 | 141.0 | 156.6 | 97.6 | 100.0 | 121.1 | 90.1 | 22.0 | 35.5 | 44.0 |
| **Peer-Reviewed** |  |  |  |  |  |  |  |  |  |  |
| Peer-reviewed | 7139 | 172.0 | 196.4 | 112.7 | 135.0 | 157.5 | 104.4 | 24.0 | 38.9 | 46.2 |
| Non-peer-reviewed | 452 | 33.0 | 57.0 | 66.8 | 9.0 | 30.2 | 53.5 | 15.0 | 26.8 | 43.3 |

G1 - Group 1, G2 - Group 2, G3 - Group 3

The results showed that articles that were submitted when COVID was endemic (Group 3) were published significantly faster than those that were submitted and published before the pandemic (Group 1), *p* < .001. The same significant results were found between these two groups for acceptance time and process time (see Table 1 for *M*s and *SD*s). Thus, these results indicate that the publication timelines were delayed specifically for those articles submitted prior to/close to the pandemic outbreak, but not for those that were submitted once COVID became endemic.

### COVID-Related Articles

When compared to non-COVID articles in general (across Groups 1-3), COVID-related articles were published significantly faster, *t* (7589) = −8.46, *p* <.001, Cohen’s *d* = 0.31. When looking into specific period, for example, among Group 3 (COVID-endemic), articles that were COVID-related had significantly shorter publication timeline (*n* = 851; *M*= 156.6; *SD* = 97.6) compared to those that were non-COVID related (*n* = 3975; *M*= 174.2; *SD* = 106.5), *t* (4824) = −4.43, *p* <.001, mean difference = −17.59, 95% *CI*: −25.36 to −9.82, Cohen’s *d* = 0.17. The same trend with significant results were also found for acceptance time and process time (see Table 1 for *M*s and *SD*s). However, non-COVID-related articles during the pandemic were still published significantly faster than those that were pre-pandemic, *t* (5977) = −8.82, *p* < .001.

### Publication Types

The NLM indexed articles as a variety of publication types (Table 2). Commentaries had the shortest publication time of 36.5 days (*n* = 271; *SD* = 37.1; *median* = 25) followed by editorials, which took on average 72.6 days (*n* = 107; *SD* = 81.3; *median* = 50). Case reports, guidelines, news, and published erratum also had relatively short timelines, but there were fewer articles representing these publication types compared to the other types. Among all types, peer-reviewed articles took significantly longer time to publish, *M*= 196.4; *SD* = 112.7, *t* (627) = 40.84, *p* < .001, mean difference = 139.38, 95% *CI*: 132.67-146.08, Cohen’s *d* = 1.26, compared to those that were not peer-reviewed (*M*= 57; *SD* = 66.8). Significant results with the same trend were found for acceptance time, and for processing time (see Table 2 and Table 1 for *M*s and *SD*s).

**Table 2.**
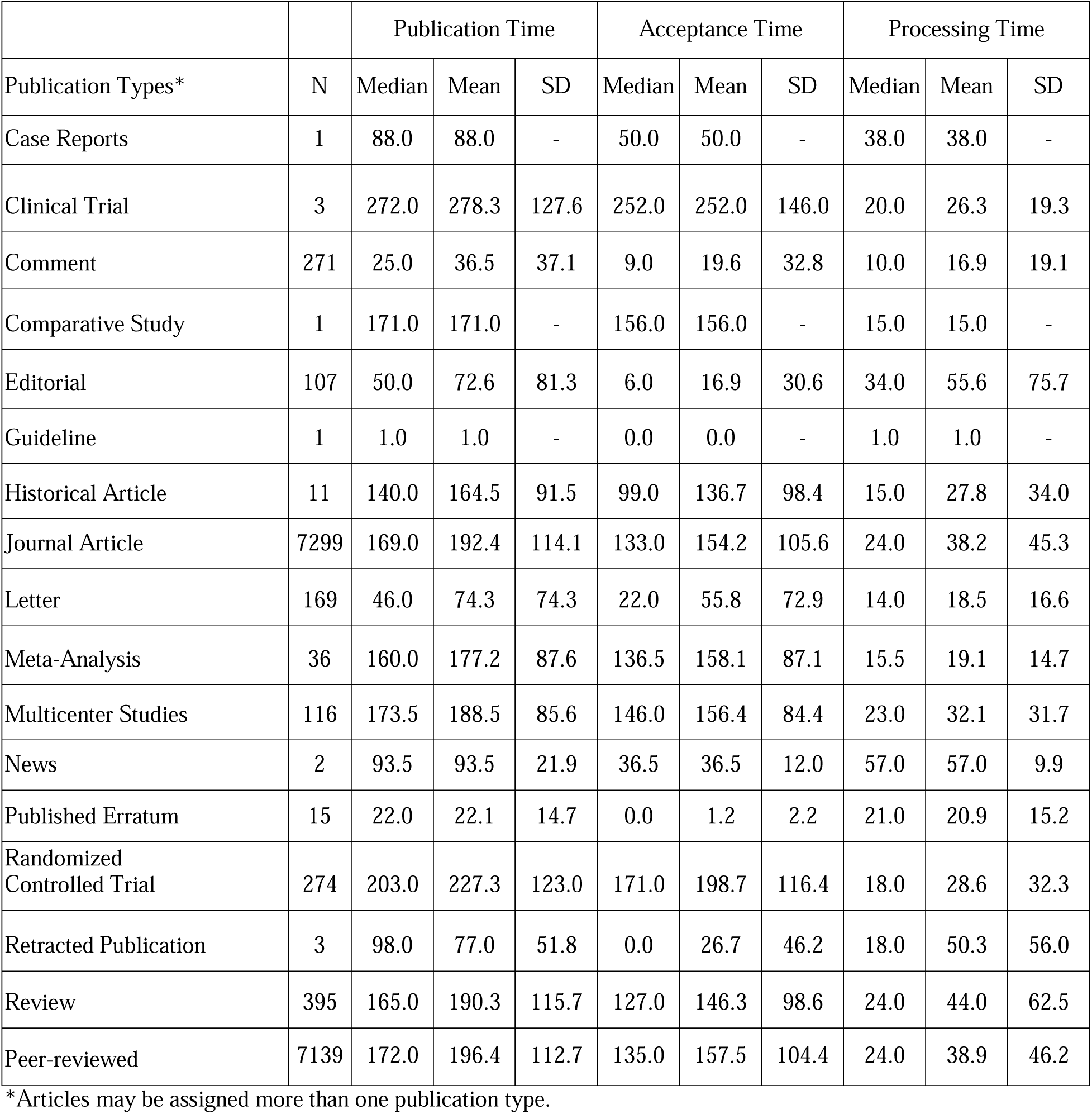
Publication timelines by publication types.

|  |  | Publication Time |  |  | Acceptance Time |  |  | Processing Time |  |  |
| --- | --- | --- | --- | --- | --- | --- | --- | --- | --- | --- |
| Publication Types* | N | Median | Mean | SD | Median | Mean | SD | Median | Mean | SD |
| Case Reports | 1 | 88.0 | 88.0 | - | 50.0 | 50.0 | - | 38.0 | 38.0 | - |
| Clinical Trial | 3 | 272.0 | 278.3 | 127.6 | 252.0 | 252.0 | 146.0 | 20.0 | 26.3 | 19.3 |
| Comment | 271 | 25.0 | 36.5 | 37.1 | 9.0 | 19.6 | 32.8 | 10.0 | 16.9 | 19.1 |
| Comparative Study | 1 | 171.0 | 171.0 | - | 156.0 | 156.0 | - | 15.0 | 15.0 | - |
| Editorial | 107 | 50.0 | 72.6 | 81.3 | 6.0 | 16.9 | 30.6 | 34.0 | 55.6 | 75.7 |
| Guideline | 1 | 1.0 | 1.0 | - | 0.0 | 0.0 | - | 1.0 | 1.0 | - |
| Historical Article | 11 | 140.0 | 164.5 | 91.5 | 99.0 | 136.7 | 98.4 | 15.0 | 27.8 | 34.0 |
| Journal Article | 7299 | 169.0 | 192.4 | 114.1 | 133.0 | 154.2 | 105.6 | 24.0 | 38.2 | 45.3 |
| Letter | 169 | 46.0 | 74.3 | 74.3 | 22.0 | 55.8 | 72.9 | 14.0 | 18.5 | 16.6 |
| Meta-Analysis | 36 | 160.0 | 177.2 | 87.6 | 136.5 | 158.1 | 87.1 | 15.5 | 19.1 | 14.7 |
| Multicenter Studies | 116 | 173.5 | 188.5 | 85.6 | 146.0 | 156.4 | 84.4 | 23.0 | 32.1 | 31.7 |
| News | 2 | 93.5 | 93.5 | 21.9 | 36.5 | 36.5 | 12.0 | 57.0 | 57.0 | 9.9 |
| Published Erratum | 15 | 22.0 | 22.1 | 14.7 | 0.0 | 1.2 | 2.2 | 21.0 | 20.9 | 15.2 |
| Randomized Controlled Trial | 274 | 203.0 | 227.3 | 123.0 | 171.0 | 198.7 | 116.4 | 18.0 | 28.6 | 32.3 |
| Retracted Publication | 3 | 98.0 | 77.0 | 51.8 | 0.0 | 26.7 | 46.2 | 18.0 | 50.3 | 56.0 |
| Review | 395 | 165.0 | 190.3 | 115.7 | 127.0 | 146.3 | 98.6 | 24.0 | 44.0 | 62.5 |
| Peer-reviewed | 7139 | 172.0 | 196.4 | 112.7 | 135.0 | 157.5 | 104.4 | 24.0 | 38.9 | 46.2 |

\*Articles may be assigned more than one publication type.

### Publications by Journal

Publication timelines for each included journal are reported in Table 3. There is variation in terms of publication timeline for each specific journal. As a robustness check for our main ANOVA analyses, we removed the journal with the longest publication time, *Anatomical Science Education*, and the results remain the same.

**Table 3.**
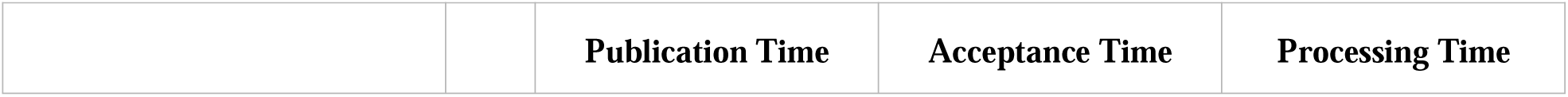
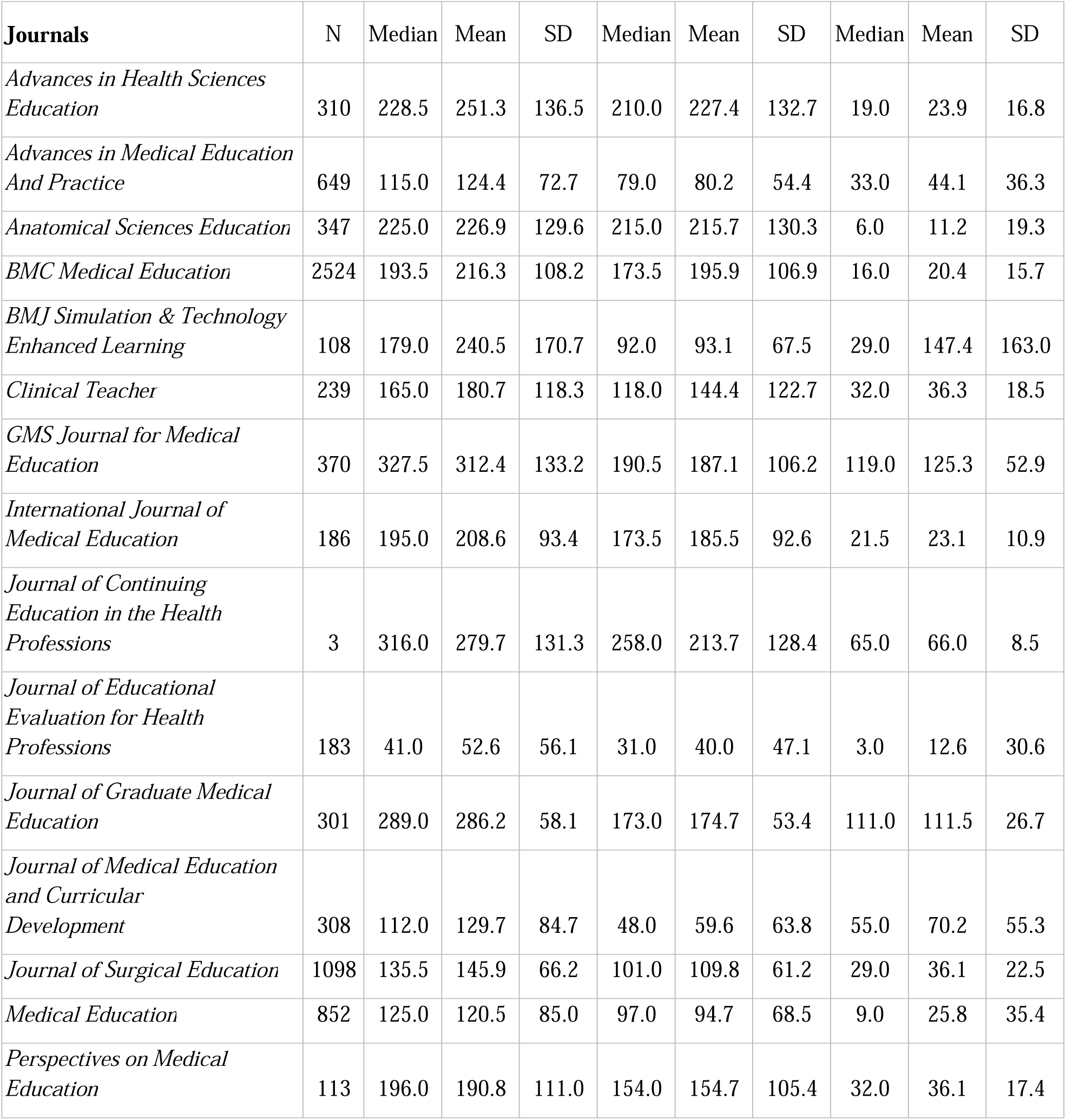
Publication timelines by journal.

|  |  | Publication Time | Acceptance Time | Processing Time |
| --- | --- | --- | --- | --- |

| **Journals** | **N** | **Median** | **Mean** | **SD** | **Median** | **Mean** | **SD** | **Median** | **Mean** | **SD** |
| --- | --- | --- | --- | --- | --- | --- | --- | --- | --- | --- |
| *Advances in Health Sciences Education* | 310 | 228.5 | 251.3 | 136.5 | 210.0 | 227.4 | 132.7 | 19.0 | 23.9 | 16.8 |
| *Advances in Medical Education And Practice* | 649 | 115.0 | 124.4 | 72.7 | 79.0 | 80.2 | 54.4 | 33.0 | 44.1 | 36.3 |
| *Anatomical Sciences Education* | 347 | 225.0 | 226.9 | 129.6 | 215.0 | 215.7 | 130.3 | 6.0 | 11.2 | 19.3 |
| *BMC Medical Education* | 2524 | 193.5 | 216.3 | 108.2 | 173.5 | 195.9 | 106.9 | 16.0 | 20.4 | 15.7 |
| *BMJ Simulation & Technology Enhanced Learning* | 108 | 179.0 | 240.5 | 170.7 | 92.0 | 93.1 | 67.5 | 29.0 | 147.4 | 163.0 |
| *Clinical Teacher* | 239 | 165.0 | 180.7 | 118.3 | 118.0 | 144.4 | 122.7 | 32.0 | 36.3 | 18.5 |
| *GMS Journal for Medical Education* | 370 | 327.5 | 312.4 | 133.2 | 190.5 | 187.1 | 106.2 | 119.0 | 125.3 | 52.9 |
| *International Journal of Medical Education* | 186 | 195.0 | 208.6 | 93.4 | 173.5 | 185.5 | 92.6 | 21.5 | 23.1 | 10.9 |
| *Journal of Continuing Education in the Health Professions* | 3 | 316.0 | 279.7 | 131.3 | 258.0 | 213.7 | 128.4 | 65.0 | 66.0 | 8.5 |
| *Journal of Educational Evaluation for Health Professions* | 183 | 41.0 | 52.6 | 56.1 | 31.0 | 40.0 | 47.1 | 3.0 | 12.6 | 30.6 |
| *Journal of Graduate Medical Education* | 301 | 289.0 | 286.2 | 58.1 | 173.0 | 174.7 | 53.4 | 111.0 | 111.5 | 26.7 |
| *Journal of Medical Education and Curricular Development* | 308 | 112.0 | 129.7 | 84.7 | 48.0 | 59.6 | 63.8 | 55.0 | 70.2 | 55.3 |
| *Journal of Surgical Education* | 1098 | 135.5 | 145.9 | 66.2 | 101.0 | 109.8 | 61.2 | 29.0 | 36.1 | 22.5 |
| *Medical Education* | 852 | 125.0 | 120.5 | 85.0 | 97.0 | 94.7 | 68.5 | 9.0 | 25.8 | 35.4 |
| *Perspectives on Medical Education* | 113 | 196.0 | 190.8 | 111.0 | 154.0 | 154.7 | 105.4 | 32.0 | 36.1 | 17.4 |

## Discussion

The pandemic phase of COVID-19 is over; yet this short-lived period had a profound impact on science and academic publishing [34]. Our findings suggest that this effect was also felt by those publishing in medical education, but these effects can be considered both positive and negative. For example, articles submitted during the COVID-overlap period took almost a year and a half to be published, which may have had consequences for individuals and the currency of the field’s knowledge base. However, in the COVID-endemic period, we observed that even with the increased number of manuscripts submitted to and published by medical education journals, the timeline for publication was significantly shorter, suggesting that there may be efficiencies worth exploring and possibly retaining into the future.

This study confirms findings from the broader scientific literature (e.g., biomedicine[35]): we found that the publication timeline for COVID-19 topics was quicker than publication timelines for non-COVID topics. This effect may have been the result of the various mechanisms that journals implemented. For example, it was found that the peer review process could be made faster because editors collaborated with editors at other journals [36] or removed hurdles such as requests for additional data [17]. While these mechanisms were introduced in a time of crisis and for a specific topic, it is important for editors and publishers to consider which, if any, should be retained into the future. Additionally, researchers should study the costs that these mechanisms might bring to the community (e.g., stress on peer reviewer effort, lessening of article quality) and the degree to which they are tolerable.

This study extends the existing literature by providing updated publication timelines and evidence-based expectations for academic productivity. In examining the COVID-pandemic effect, articles submitted between January 1, 2018 and March 10, 2020, and those published after March 10, 2020, took substantially longer to move through the publication process. So, while many universities granted “the gift of time” by automatically extending faculty tenure clocks during the pandemic [37], tenure and promotion committees must consider that many articles were impacted, including not just those that were submitted during the pandemic phase, but also articles under review or in revisions prior to COVID that were significantly affected. Thus, faculty hired between 2018 and 2020 might be expected to have less research productivity (at least in terms of published articles). Faculty seeking tenure and promotion could use these findings in a statement about how COVID shaped their academic productivity. Lastly, we observed that the article type (e.g., commentary, review article) should also be taken into consideration when assessing productivity, as time to publication can vary considerably between publication types.

Previous research found that medical education articles took an average of 263 days to publish between 2008 and 2018 [1]. Since March 11, 2020, articles are taking an average of 226 days from submission to appearing in PubMed. While shorter, this more recent timeline stands in contrast to the estimated 100 days it takes for the biomedical articles to be published [24], which has been criticized as a lengthy publication delay with potential negative effects on scientific progress [38]. As a field, medical education must consider what such a delay means for the timeliness of the evidence that is used to make decisions and craft educational policies.

### Limitations and Areas for Future Research

This study has limitations. First, only 15 of the MEJ-24 journals supplied publication timeline data. However, this represented a sampling frame of over half (66.6%) of the articles published in the study’s timeframe. It is possible that journals which did not report publication timeline data in their metadata represent a form of non-response bias, if they are systematically different from the journals that did report timelines in their metadata. However, in comparison to Maggio et. al’s earlier study [1], our current sample represents an increase in availability of publication metadata. Second, we relied upon NLM indexers to determine whether or not an article was about COVID. While information scientists designed and vetted the search approach, it is possible that some articles were inadvertently missed. This study focused on the effects that the COVID pandemic may have had on publication timelines in medical education. However, it is worth remembering that many other events occurred during this time period that also could have influenced publication timelines, including several social and environmental events like the Black Lives Matter movement, Hong Kong protests, and Australian bushfires.

The quantitative and descriptive approach used in this study leaves unanswered many questions about publication timelines in medical education. For instance, publication timelines are composed of multiple steps undertaken by a variety of individuals (e.g., authors, editors, reviewers, copy editors, type setters); however, the data analyzed here provides limited insight into how each of these individuals contributed to the variable publication timelines observed. In terms of impact, it is unclear if and how substantive publication delays in the COVID-overlap group affected early career authors. Additionally, it is unknown whether the steps journal editors took actually alleviated the problems associated with reviewer responsiveness or promoted COVID-specific scholarship. In light of these limitations, future qualitative studies should explore the lived experiences of authors and editors to better understand the ways in which COVID may have shaped medical education publishing.

Conflicts of interest: None reported

Funding/Support: No specific funding was received for this work

Other disclosures: None reported

Ethical approval: Not applicable

Disclosures: None reported

Disclaimer: The views expressed in this article are those of the authors and do not necessarily reflect the official policy or position of the Uniformed Services University of the Health Sciences, Henry M. Jackson Foundation, the Department of Defense, or the U.S. Government.

Previous presentations: None

Data: Costello JA, Maggio LA, Brown KR, Artino Jr AR, Durning S, Ma T. Revisiting the time to publication in medical education: An analysis of publication timelines between 2019-2022 [Data set]. Zenodo. Published 2023. Available at: https://doi.org/10.5281/zenodo.10433647

## Data Availability

All data produced are available online at: Costello JA, Maggio LA, Brown KR, Artino Jr AR, Durning S, Ma T. Revisiting the time to publication in medical education: An analysis of publication timelines between 2019-2022 [Data set]. Zenodo. Published 2023. Available at: https://doi.org/10.5281/zenodo.10433647

---

**Appendix A.** MEJ-24 Journals with Publication Timeline Data Available for Analysis.

| Journals | Articles published | Publication Timeline Data Available | Publication Timeline Data Meeting Inclusion Criteria** | Avg. time to publication (SD) | Avg. time to acceptance (SD) | Average processing time (SD) |
| --- | --- | --- | --- | --- | --- | --- |
| *Academic Medicine* | 2980 |  |  |  |  |  |
| *Advances in Health Sciences Education* | 380 | 356 | 310 | 251.3 (136.48) | 227.4 (132.7) | 23.9 (16.8) |
| *Advances in Medical Education And Practice* | 822 | 650 | 649 | 124.4 (72.7) | 80.2 (54.4) | 44.1 (36.3) |
| *African Journal of Health Professions Education* | 4 |  |  |  |  |  |
| *Anatomical Sciences Education* | 400 | 389 | 347 | 226.9 (129.6) | 215.7 (130.3) | 11.2 (19.3) |
| *BMC Medical Education* | 2850 | 2776 | 2524 | 216.3 (108.2) | 195.9 (106.9) | 20.4 (15.7) |
| *BMJ Simulation & Technology Enhanced Learning* | 343 | 333 | 108 | 240.6 (170.7) | 93.1 (67.5) | 147.4 (163) |
| *Canadian Medical Education Journal\** |  |  |  |  |  |  |
| *Clinical Teacher* | 652 | 245 | 239 | 180.7 (118.3) | 144.4 (122.7) | 36.3 (18.5) |
| *Education for Health* | 158 |  |  |  |  |  |
| *Focus on Health Professional Education\** |  |  |  |  |  |  |
| *GMS Journal for Medical Education* | 414 | 414 | 370 | 312.4 (133.2) | 187.1 (106.2) | 125.3 (52.9) |
| *International Journal of Medical Education* | 224 | 224 | 186 | 208.6 (93.4) | 185.5 (92.6) | 23.1 (10.9) |
| *Journal of Continuing Education in the Health Professions* | 269 | 3 | 3 | 279.7 (131.3) | 213.7 (128.4) | 66 (8.5) |
| *Journal of Educational Evaluation for Health Professions* | 192 | 192 | 183 | 52.6 (56.1) | 40 (47.1) | 12.6 (30.6) |
| *Journal of Graduate Medical Education* | 926 | 346 | 301 | 286.2 (58.1) | 174.7 (53.4) | 111.5 (26.7) |
| *Journal of Medical Education and Curricular Development* | 378 | 354 | 308 | 129.7 (84.7) | 59.6 (63.8) | 70.2 (55.3) |
| *Journal of Surgical Education* | 1201 | 1190 | 1098 | 145.9 (66.2) | 109.8 (61.2) | 36.1 (22.5) |
| *Medical Education* | 1354 | 913 | 852 | 120.5 (85) | 94.7 (68.5) | 25.8 (35.4) |
| *Medical Education Online* | 405 |  |  |  |  |  |
| *Medical Teacher* | 1556 |  |  |  |  |  |
| *Perspectives on Medical Education* | 315 | 116 | 113 | 190.8 (111) | 154.7 (105.4) | 36.1 (17.4) |
| *Simulation in Healthcare* | 411 |  |  |  |  |  |
| *Teaching and Learning in Medicine* | 310 |  |  |  |  |  |
| Total | 16544 | 8501 | 7591 |  |  |  |

\*Not indexed in PubMed
\*\* Submitted for peer-review on or after January 1, 2018 and Published by December 31, 2022

## References

1. Maggio LA, Bynum WE 4th, Schreiber-Gregory DN, Durning SJ, Artino AR Jr. When will I get my paper back? A replication study of publication timelines for health professions education research. Perspect Med Educ. 2020;9(3):139–146. DOI:10.1007/s40037-020-00576-2

2. Raynaud M, Goutaudier V, Louis K, et al. Impact of the COVID-19 pandemic on publication dynamics and non-COVID-19 research production. BMC Med Res Methodol. 2021;21(1):255. DOI:10.1186/s12874-021-01404-9

3. Squazzoni F, Bravo G, Grimaldo F, García-Costa D, Farjam M, Mehmani B. Gender gap in journal submissions and peer review during the first wave of the COVID-19 pandemic. A study on 2329 Elsevier journals. PLoS One. 2021;16(10):e0257919. DOI:10.1371/journal.pone.0257919

4. Riccaboni M, Verginer L. The impact of the COVID-19 pandemic on scientific research in the life sciences. PLoS One. 2022;17(2):e0263001. DOI:10.1371/journal.pone.0263001

5. Clark J. How covid-19 bolstered an already perverse publishing system. BMJ. 2023;380:689. DOI:10.1136/bmj.p689

6. Vaux E, Moss F. Covid-19 – A Game Changer for Medical Education? Reflections from England. IJQHC Communications. 2021;lyab017. DOI:10.1093/ijcoms/lyab017

7. Gayet-Ageron A, Ben Messaoud K, Richards M, Schroter S. Female authorship of covid-19 research in manuscripts submitted to 11 biomedical journals: cross sectional study. BMJ. 2021;375:n2288. DOI:10.1136/bmj.n2288

8. Horbach S. Pandemic publishing: Medical journals strongly speed up their publication process for COVID-19. Quantitative Science Studies. 2020;1(3):1056–1067. DOI:10.1162/qss_a_00076

9. Shan J, Ballard D, Vinson DR. Publication Non Grata: The Challenge of Publishing Non-COVID-19 Research in the COVID Era. Cureus. 2020;12(11):e11403. DOI:10.7759/cureus.11403

10. Aviv-Reuven S, Rosenfeld A. Publication patterns’ changes due to the COVID-19 pandemic: a longitudinal and short-term scientometric analysis. Scientometrics. 2021;126(8):6761–6784. DOI:10.1007/s11192-021-04059-x

11. Gabster BP, van Daalen K, Dhatt R, Barry M. Challenges for the female academic during the COVID-19 pandemic. Lancet. 2020;395(10242):1968–1970. DOI:10.1016/S0140-6736(20)31412-4

12. Kwan JM, Noch E, Qiu Y, et al. The Impact of COVID-19 on Physician-Scientist Trainees and Faculty in the United States: A National Survey. Acad Med. 2022;97(10):1536–1545. DOI:10.1097/ACM.0000000000004802

13. Deryugina T, Shurchkov O, Stearns J. COVID-19 disruptions disproportionately affect female academics. AEA Papers and Proceedings. 2021;111:164–168. DOI: 10.1257/pandp.20211017

14. Krukowski RA, Jagsi R, Cardel MI. Academic Productivity Differences by Gender and Child Age in Science, Technology, Engineering, Mathematics, and Medicine Faculty During the COVID-19 Pandemic. J Womens Health (Larchmt). 2021;30(3):341–347. DOI:10.1089/jwh.2020.8710

15. Bender S, Brown KS, Hensley Kasitz DL, Vega O. Academic women and their children: Parenting during COVIDLJ19 and the impact on scholarly productivity. Family Relations. 2022;71(1):46–67. DOI:10.1111/fare.12632

16. Hindawi Ltd. C19 Rapid Review Initiative expands to include 20 publishers and organizations. EurekAlert! From the American Association for the Advancement of Science (AAAS). https://www.eurekalert.org/news-releases/844794 (accessed 30 October 2023).

17. Eisen MB, Akhmanova A, Behrens TE, Weigel D. Publishing in the time of COVID-19. Elife. 2020;9:e57162. DOI:10.7554/eLife.57162

18. Advances in Health Sciences Education. COVID-19 and impact on peer review. https://www.springer.com/journal/10459/updates/17818222. (accessed 30 October 2023).

19. Sevryugina YV, Dicks AJ. Publication practices during the COVID-19 pandemic: Expedited publishing or simply an early bird effect?. Learn Publ. Published online June 30, 2022. DOI:10.1002/leap.1483

20. Palayew A, Norgaard O, Safreed-Harmon K, Andersen TH, Rasmussen LN, Lazarus JV. Pandemic publishing poses a new COVID-19 challenge. Nat Hum Behav. 2020;4(7):666–669. DOI:10.1038/s41562-020-0911-0

21. Schimanski LA, Alperin JP. The evaluation of scholarship in academic promotion and tenure processes: Past, present, and future. F1000Res. 2018;7:1605. DOI:10.12688/f1000research.16493.1

22. Rice DB, Raffoul H, Ioannidis JPA, Moher D. Academic criteria for promotion and tenure in biomedical sciences faculties: cross sectional analysis of international sample of universities. BMJ. 2020;369:m2081. DOI:10.1136/bmj.m2081

23. Sackett DL, Rosenberg WM, Gray JA, Haynes RB, Richardson WS. Evidence based medicine: what it is and what it isn’t. BMJ. 1996;312(7023):71–72. DOI: 10.1136/bmj.312.7023.71.

24. Powell K. Does it take too long to publish research?. Nature. 2016;530(7589):148–151. DOI:10.1038/530148a

25. Maggio LA, Ninkov A, Frank JR, Costello JA, Artino AR Jr. Delineating the field of medical education: Bibliometric research approach(es). Med Educ. 2022;56(4):387–394. DOI:10.1111/medu.14677

26. World Health Organization. WHO Director - General’s opening remarks at the media briefing on COVID-19 - 11 March 2020. https://www.who.int/director-general/speeches/detail/who-director-general-s-opening-remarks-at-the-media-briefing-on-COVID-19 11-march-2020. (accessed 30 October 2023).

27. Himmelstein D. The history of publishing delays. Satoshi Village: the blog of Daniel Himmelstein. 2016. https://blog.dhimmel.com/history-of-delays/. (accessed 21 August 2023).

28. Himmelstein D. The history of publishing delays. https://github.com/dhimmel/delays/blob/master/README.md. (accessed 21 August 2023).

29. Kans J. Entrez Direct: E-utilities on the Unix Command Line. 2013 Apr 23 [Updated 2023 Aug 22]. In: Entrez Programming Utilities Help [Internet]. Bethesda (MD): National Center for Biotechnology Information (US); 2010-. https://www.ncbi.nlm.nih.gov/books/NBK179288/ (accessed 30 October 2023).

30. Chen Q, Allot A, Lu Z. Keep up with the latest coronavirus research. Nature. 2020;579(7798):193. DOI:10.1038/d41586-020-00694-1

31. Chen Q, Allot A, Lu Z. LitCovid: an open database of COVID-19 literature. Nucleic Acids Res. 2021;49(D1):D1534–D1540. DOI:10.1093/nar/gkaa952

32. SPSS version 28 (IBM SPSS Statistics, Chicago, IL, 2021).

33. Costello JA, Maggio LA, Brown KR, Artino Jr AR, Durning S, Ma T. Revisiting the time to publication in medical education: An analysis of publication timelines between 2019-2022 [Data set]. Zenodo. DOI:10.5281/zenodo.10433647

34. Lucey CR, Davis JA, Green MM. We Have No Choice but to Transform: The Future of Medical Education After the COVID-19 Pandemic. Acad Med. 2022;97(3S):S71–S81. DOI:10.1097/ACM.0000000000004526

35. Ioannidis JPA, Bendavid E, Salholz-Hillel M, Boyack KW, Baas J. Massive covidization of research citations and the citation elite. Proc Natl Acad Sci U S A. 2022;119(28):e2204074119. DOI:10.1073/pnas.2204074119

36. Putman MS, Ruderman EM, Niforatos JD. Publication Rate and Journal Review Time of COVID-19-Related Research. Mayo Clin Proc. 2020;95(10):2290–2291. DOI:10.1016/j.mayocp.2020.08.017

37. Smith JL, Vidler LL, Moses MS. The "Gift" of Time: Documenting Faculty Decisions to Stop the Tenure Clock During a Pandemic. Innov High Educ. 2022;47(5):875–893. DOI:10.1007/s10755-022-09603-y

38. Palese A, Coletti S, Dante A. Publication efficiency among the higher impact factor nursing journals in 2009: a retrospective analysis. Int J Nurs Stud. 2013;50(4):543–551. DOI:10.1016/j.ijnurstu.2012.08.019

